# Worldwide and Regional Forecasting of Coronavirus (Covid-19) Spread using a Deep Learning Model

**DOI:** 10.1101/2020.05.23.20111039

**Authors:** Cem Direkoglu, Melike Sah

## Abstract

In December 2019, Covid-19 epidemic was identified in Wuhan, China. Covid-19 may cause fatality especially among elderly, and people with chronic health problems. After human to human transmissions of the disease, it has rapidly spread throughout China, and then the outbreak has reached to neighboring countries in Asia. Now, the spread of the virus is accelerating in the world, and increasing number of new cases has been reported daily in Europe, Middle East, Africa and America regions. Recently, World Health Organization (WHO) also announced Covid-19 as a Pandemic. As of 3 April, worldwide around more than 1 million cases and around 60,000 fatalities are reported. Thus, forecasting regional and worldwide outbreak size of Covid-19 is important in order to take necessary actions regarding to preparedness plans and mitigation interventions. In this work, we design a deep learning model, which is an effective artificial intelligence method, to provide regional and worldwide forecasts. Particularly for worldwide, our approach predicts the cumulative number of cases, cumulative number of deaths and daily new cases. For Europe and Middle East regions, we predict the cumulative number of cases, and for Mainland China we predict daily new cases and the cumulative number of deaths. We predict the next 10 days based on the previously reported actual time series data of Covid-19. For worldwide forecasts, we use the data provided by Worldometers. For Europe and Middle East forecasts, we use the data provided by World Health Organization, and for China Mainland forecasts, the data is obtained from Chinese Centre for Disease Control and Prevention. This is the first time that a deep learning model has been employed for Covid-19 spread prediction, solely based on the known reported cases of Covid-19. The proposed deep learning architecture consists of Long Short Term Memory (LSTM) layer, dropout layer, and fully connected layers to predict regional and worldwide forecasts. We evaluate our approach with Root Mean Square Error (RMSE) metric. For forecasting, we use the network models that give the minimum RMSE on the last 3 days of actual data. Networks, which achieves the minimum RMSE on the last 3 days, are used to predict the next 10 days. Every day, the spread and situations are changing. Our approach can take into account these realtime changes; the deep learning model can be re-trained with new daily data and perform real-time forecasting. Results show that the proposed deep learning model is promising, it can predict possible scenarios regionally and globally for the spread of Covid-19.

## 1. Introduction

World heard about the dangerous disease originated in Wuhan, China, by the end of December 2019. Later, it is identified as a novel type of coronavirus and named as Covid-19 epidemic [1]. The epicenter and the worst affected region by Covid-19 is Hubei province of China, where most of the reported cases and human tragedies were initially from there. Covid-19 is very contagious; meaning that the disease transmissibility (reproductive number, R_0_) is high. A single infected person will transmit the virus with a R_0_ of 1.4 to 2.5 according to World Health Organization [2]. Other studies estimate the R_0_ between 2.24 and 3.58 [1], between 2.47 and 2.86 [12], and between 3.30 and 5.47 [3]. These figures suggest that Covid-19 can spread more rapidly if it is not contained. Unfortunately, Covid-19 does not have any known cure or vaccine yet, it has long incubation period, as well as it may cause pneumonia that may lead to deaths especially among elderly and people with chronic health problems like diabetics, hypertension and cancer. Thus, predicting the spread and taking necessary prevention measures and implementations is vital.

Recently, outbreak of novel coronavirus is slowing down in China after major measures taken by authorities [4]. However, the outbreak is accelerating in the World particularly in Europe, Middle East, America and Asia (except China) regions [5][6]. Many countries, such as Italy, Spain, Iran and United States, are affected very badly nowadays by the virus. Recently, World Health Organization (WHO) also announced Covid-19 as a Pandemic. Therefore, it is vital to forecast regional and worldwide outbreak size of Covid-19 in order to take necessary actions regarding to preparedness plans and mitigation interventions. Especially, there is a need to predict the number of daily and cumulative cases and deaths to take necessary actions.

In early days of Covid-19, most of the researchers focused on predicting the transmissibility (reproduction number, R_0_) of the novel coronavirus. One of the first studies was carried out by Zhao et al [3] in early stages. They use the epidemic curve of the time series data, and estimate R_0_ value between 3.30 and 5.47. Hermanowicz [7] use a simple logistic growth model to estimate growth rate in China. Li [8] uses a stochastic method for predicting transmission potential. On the other hand, some researchers estimate the cumulative reported cases based on data analysis. Roosa et al. provides short-terms forecasts of reported cases in China [9], and in Guangdong and Zhejiang [10] based on three different models namely generalized logistic growth model, Richards growth model and a sub-epidemic wave model, respectively. Some studies investigate the international spread of Covid-19 between countries. Ivorra and Ramos [11] apply Between Countries Disease Spread (Be-CoDiS), which is a spatio-temporal epidemiological model for studying transmission of diseases between and within countries. Wu et al. [12] uses monthly flight bookings and mobility information about individuals to estimate the outbreak of Covid-19 in China and from China to other countries. However, only few studies forecast the growth size of the Covid-19 epidemic. Batista [13] applies logistic growth regression model to predict the size of the outbreak worldwide. Hu et al. [14] uses a modified stacked auto-encoder model which is a traditional neural network (not deep) for predicting the size, length and ending-time of Covid-19 in China. Most of the methods presented above are statistical methods. Mathematical modelling based on dynamical equations can also provide information about pandemic dynamics. Such as a susceptible-exposed-infected-removed (SEIR) model [26] that is based on traditional susceptible-infected-removed (SIR) model has been introduced to predict the Covid-19 outbreak in Spain.

In this work, we rather use a different strategy and apply a deep learning model on the reported cases of Covid-19. We design a deep neural network, which consist of Long Short Term Memory (LSTM) layer, dropout layer, and fully connected layers, to analyze the reported Covid-19 cases and predict the possible future scenarios for the spread in China, Europe, Middle East and worldwide. Especially for worldwide, our approach predicts the cumulative number of cases, cumulative number of deaths and daily new cases. For Europe and Middle East regions, we predict the cumulative number of cases, and for Mainland China, we predict daily new cases and the cumulative number of deaths. All predictions are done for the next 10 days given the actual time series data of Covid-19. This is the first time that a deep learning based solution is proposed for Covid-19 spread prediction solely based on the known reported cases of Covid-19. LSTM layer is a type of Recurrent Neural Network (RNN) that utilizes learning from concurrent data. We evaluate our approach on the last 3 days of actual data using Root Mean Square Error (RMSE) metric. We present results from the networks that give the minimum RMSE values. Thus, the network with minimum RMSE is used to predict the future scenarios. As new data arise daily, the network can be re-train in order to adjust the real-time predictions. These short-term forecasts are important to take the necessary interventions to alleviate a pandemic. Next, we present the deep neural network used for forecasting. Then, we evaluate our approach with RMSE, and finally illustrate and discuss the possible scenarios for Covid-19 spread regionally and worldwide. The proposed approach is promising for Covid-19 spread prediction.

## 2. Deep Learning with Long Short-Term Memory (LSTM) Network

Deep learning is a subarea of artificial intelligence that tries to model how the human brain processes data and makes decisions [15][16][17]. Deep learning has been applied to many fields including computer vision, speech recognition, natural language processing, social network filtering, machine translation, bioinformatics, drug design, and medical image analysis. Popular deep learning architectures are known as deep feed-forward neural networks, deep belief networks, recurrent neural networks and convolutional neural networks. Deep learning proved popular and very effective because it has produced results comparable to and in some cases surpassing human expert performance [18][19][20].

A powerful type of deep neural network designed to handle sequence dependence is the recurrent neural networks (RNNs). Unlike standard feedforward neural networks, RNNs have feedback connections [15]. The Long Short-Term Memory (LSTM) network is an advanced version of recurrent neural network [21]. The LSTM network was developed to deal with the exploding and vanishing gradient problems encountered during the training of traditional RNNs, as well as to solve the long-term sequence dependence problem of RNNs. Deep learning architecture consist of LSTM layer (network) can address difficult sequence problems in deep learning and achieves state-of-the-art results in time series prediction [22].

A LSTM unit (Figure 1) consist of a cell, which contains a forget gate (f_t_), an input gate (i_t_) and an output gate (o_t_) at time step *t* as shown in Figure 1. Using these gates, a LSTM cell models the flow of information over consecutive time steps. An input gate conditionally decides which values from the input to update the memory state. Output gate conditionally decides what to output based on input and the memory block. Forget gate conditionally decides what data to remove from the block. Input gate, output gate and forget gate regulate the flow of information in the cell. Each gate uses the sigmoid activation functions to control whether they are triggered or not, making the change of state and addition of information flowing through the block conditional as shown in Figure 1. In Figure 1, *C*, *x, h* represent cell, input and output values. Subscript *t* denotes time step value, i.e., *t−1* is the previous state (or from time *t*−1) and *t* denotes the current state values. The symbol *S* is the sigmoid function and *tanh* is the hyperbolic tangent function. Operator + is the element-wise summation and *x* is the element-wise multiplication. The computations of the gates are described in the equations below [23], [24].

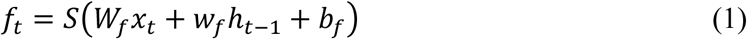

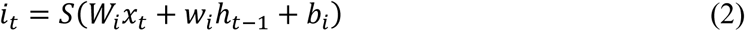

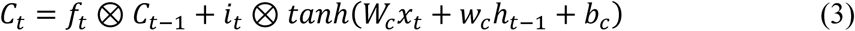

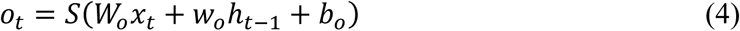

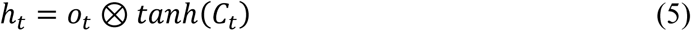

where *f, i, o* are the forget, input and output gate vectors respectively. *W, w*, *b* and ⊗ represent weights of input, weights of recurrent output, bias and element-wise multiplication respectively.

**Figure 1:**
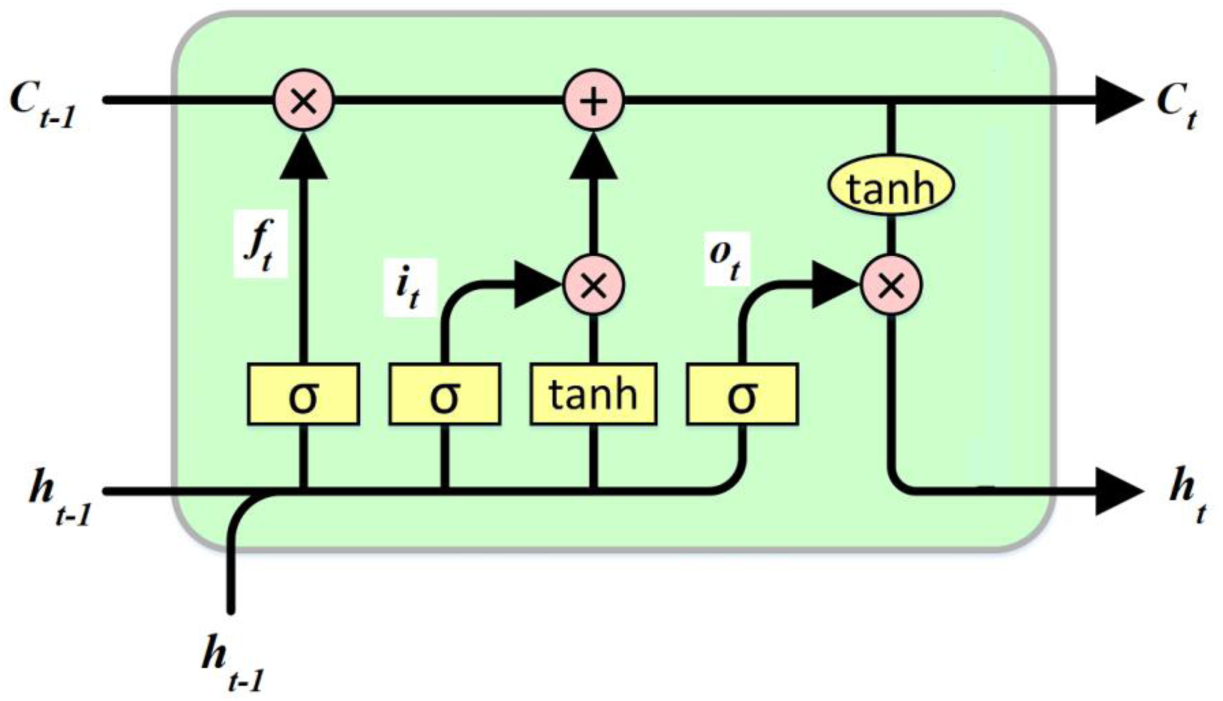
The LSTM unit

## 3. Deep Learning Architecture and Training for Covid-19 Forecasting

Convid-19 spread forecasting was experimented with different deep learning architectures consisting of varying number of LSTM, dropout and fullyconnected layers. We observe that an optimal deep learning architecture for Covid-19 spread prediction can be the one that consist of one LSTM layer, one dropout layer, and three fully connected layers as shown in Figure 2. In this architecture, shown in Figure 2, the input is a time series sequence that represents the previously reported daily cases. The LSTM layer consists of 200 units. After the LSTM layer, we have a dropout layer to prevent the model from overfitting. The dropout layer randomly set input elements to zero with probability *p* = 0.2 during the training phase. Then, we form two cascaded fully connected layer, each has 100 units, and finally one more fully connected layer with one unit, and the last layer is regression output layer. The input sequence is used to predict future values at the regression layer.

**Figure 2:**
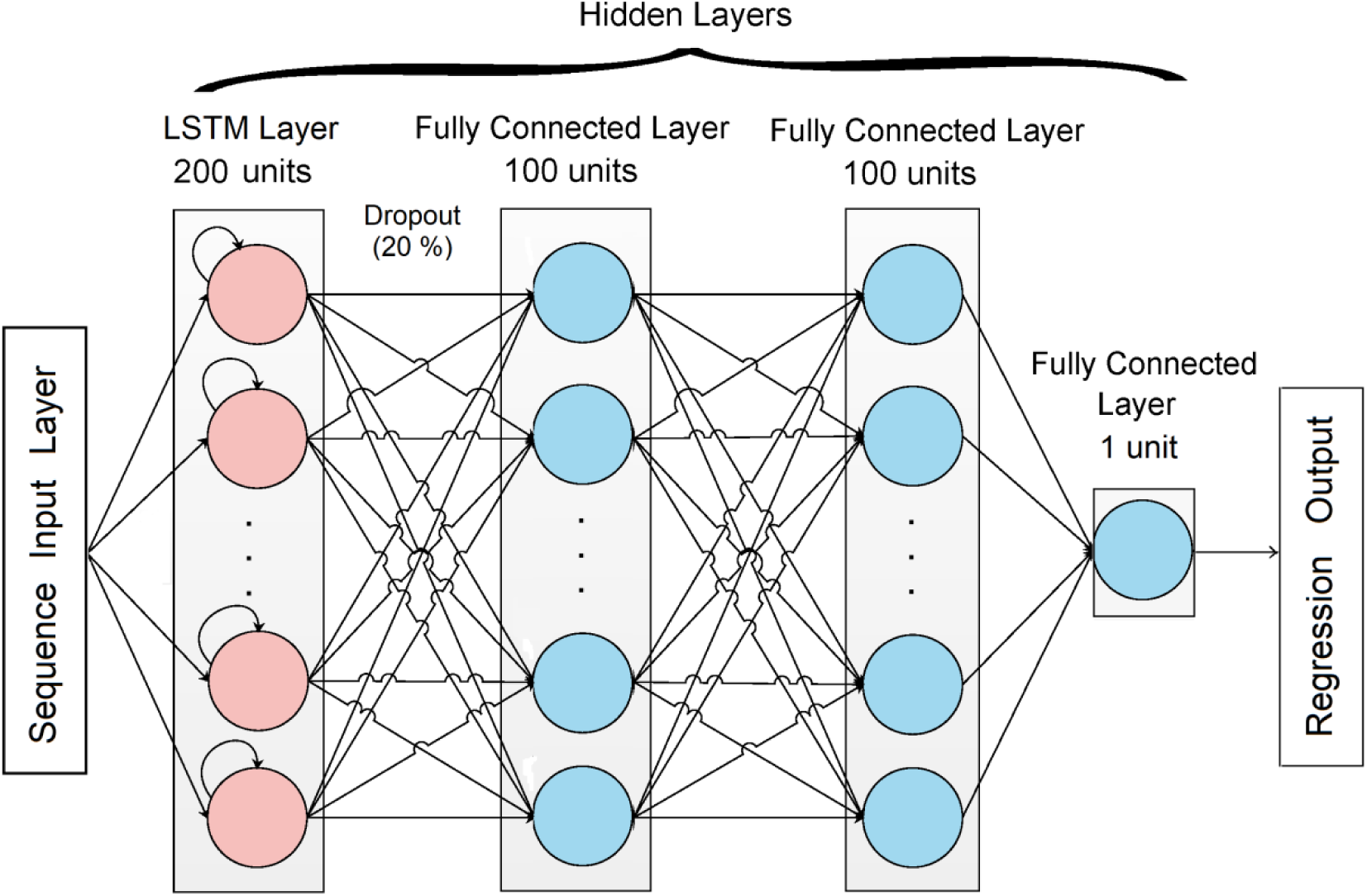
The deep learning architecture used for Covid-19 prediction

During the training, an adaptive moment estimation (ADAM) method [25] is utilized as a solver. ADAM is a method for stochastic optimization. The learning rate is 0.05, gradient decay factor 0.9, squared gradient decay factor 0.999, gradient threshold method is L2 norm, learning rate schedule is piecewise, mini batch size is 10, and the maximum number of epochs is 250. These parameter values are determined experimentally to achieve the best performances (RMSE values) with the network. The same network structure and parameter values are utilized for all forecasts of Covid-19 regionally and worldwide. Table 1 also shows the details of the network such as number of activations at each layer, total learnable parameters at each layer, and in the whole network.

**Table 1:** Parameter analysis of the deep learning architecture

| Layer | Name | Activations | Learnables | Total Learnables |
| --- | --- | --- | --- | --- |
| 1 | Sequence input (1-Dimension) | 1 | - | 0 |
| 2 | LSTM | 200 | Input Weights: 800x1<br>Recurrent Weights: 800x200<br>Bias: 800x1 | 161600 |
| 3 | Dropout (20 %) | 200 | - | 0 |
| 4 | Fully Connected | 100 | Weights: 100x200<br>Bias: 100x1 | 20100 |
| 5 | Fully Connected | 100 | Weights: 100x100<br>Bias: 100x1 | 10100 |
| 6 | Fully Connected | 1 | Weights: 1x100<br>Bias: 1x1 | 101 |
| 7 | Regression Output | - | - | 0 |
| **Total Learnable Parameters in the Network =** |  |  |  | **191901** |

## 4. Real-Time Forecasts of Covid-19 in China, Europe, Middle East and Worldwide

### 4.1 Data Sources and Method

Experiments are conducted on publicly available reported cases of Covid-19. The data used to support the findings of this study are included within the article [4][5][6]. For forecasts of Covid-19 in China, we use time series data of daily reports of Chinese Centre for Disease Control and Prevention from 10 January 2020 to 3 April 2020 [4]. The data includes the total number of daily new cases of Covid-19, the total number of deaths and the total number of recoveries in China. For forecasting the outbreak of Covid-19 in Europe and Middle East, we use time series data of World Health Organization situation reports from 17 January to 3 April 2020 [5]. The data includes the total number of daily new cases of Covid-19 in Europe and Middle East regions. For worldwide predictions, we use time series data of WorldoMeter coronavirus statistics [6] from 22 January 2020 to 3 April 2020. The data includes the total number of daily new cases of Covid-19 worldwide, the cumulative total number of Covid-19 cases worldwide and the total number of deaths worldwide. The size of time series data for each forecast provided below in Table 2.

**Table 2:** Training and Test Data

|  | From | To | Number of days (size of training data) Until 31rd of March | Number of days (size of test data) 1-3 April |
| --- | --- | --- | --- | --- |
| Cumulative cases of Covid-19 worldwide [6] | 22 January 2020 | 3 April 2020 | 74 | 3 |
| Total number of deaths worldwide [6] | 22 January 2020 | 3 April 2020 | 74 | 3 |
| Daily new cases worldwide [6] | 23 January 2020 | 3 April 2020 | 73 | 3 |
| Daily new cases in China [4] | 17 January 2020 | 3 April 2020 | 78 | 3 |
| Total number of deaths in China [4] | 10 January 2020 | 3 April 2020 | 85 | 3 |
| Cumulative cases of Covid-19 in Europe [5] | 25 January 2020 | 3 April 2020 | 70 | 3 |
| Cumulative cases of Covid-19 in Middle East [5] | 29 January 2020 | 3 April 2020 | 66 | 3 |

For predictions, we train the deep learning model with time series data that is the reported cases of Covid-19 up until the last three days (i.e. we use the data until 31rd of March for training and the last three days for testing). Then, we predict the last three days and compare with the actual data using Root Mean Square Error (RMSE) metric that is a commonly used error calculation metric in forecasting problems. The RMSE equation is given below,

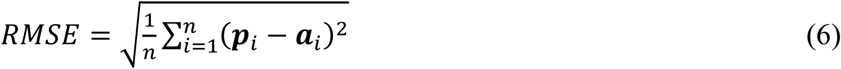

Where ***p***, ***a***, and *n* represent predicted values, actual values and the number of predictions, respectively. Since the training of the deep learning model is based on stochastic optimization [25], the same network generates different regressors after training. The trained models, which have minimum RMSE values, are used to predict the next 10 days ahead (possible future scenarios). The same strategy has been employed for all predictions. It is also important to note that, for a better fit and to prevent the training from diverging, the training data is standardized to have zero mean and unit variance as given in Equation (7), where ***t***, *μ*, and *σ* are the training vector, mean of the training vector, standard deviation of the training vector, respectively. At prediction time, we also standardize the test data using the same parameters as we used in the training data, shown in Equation (8) where ***e*** is the test vector. Finally, we unstandardize the predictions as shown in Equation (9), using the parameters calculated in Equation (7), where ***r*** is the standardized predicted vector, and ***p*** is the unstandardized predicted vector.

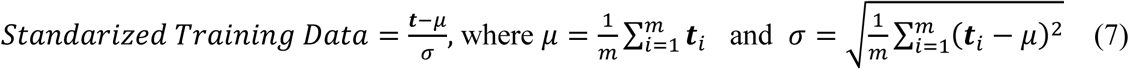

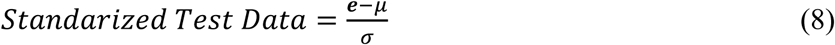

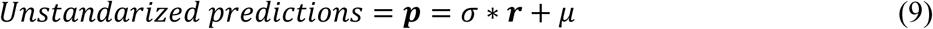

### 4.2 Results and Discussions for Worldwide Forecasts

To estimate the size of the outbreak worldwide, we predict cumulative total number of cases, total daily new cases and the possible number of deaths worldwide for the next 10 days. We present possible future scenarios with respect to observed different RMSE values on the last known three days, as we explained in the previous section.

There are two scenarios for global cumulative outbreak size: (1) As shown by Figure 3, the global outbreak size may reach to 2 million within 10 days and continue to grow linearly (RMSE=30758). (2) As shown by Figure 4, the global outbreak size may reach to 2,600,000 in 10 days and may continue to grow linearly (RMSE=39699). We also present different scenarios for daily new cases of Covid-19 worldwide. According to the deep learning model, daily new cases will increase and we do not expect a decrease in daily new cases within the next 10 days. These different scenarios are as follows: (1) Daily new cases may be in the range of 90,000 and 120,000 within the next week shown by Figure 5 (RMSE=1953.5). (2) There might be a jump in the number of reported cases within the next week, shown in Figure 6 (RMSE=64337), where daily new cases may reach to 160,000. (3) Daily new cases may even reach to 200,000 as illustrated in Figure 7 (RMSE=63850). For possible number of deaths worldwide, the deep learning approach forecasts that the number of deaths may reach to 100,000 within 10 days (RMSE=5657.1) (see Figure 8). In a worst scenario, number of deaths may reach to 150,000 within the next 10 days (RMSE=7703.3) as shown by Figure 9.

**Figure 3:**
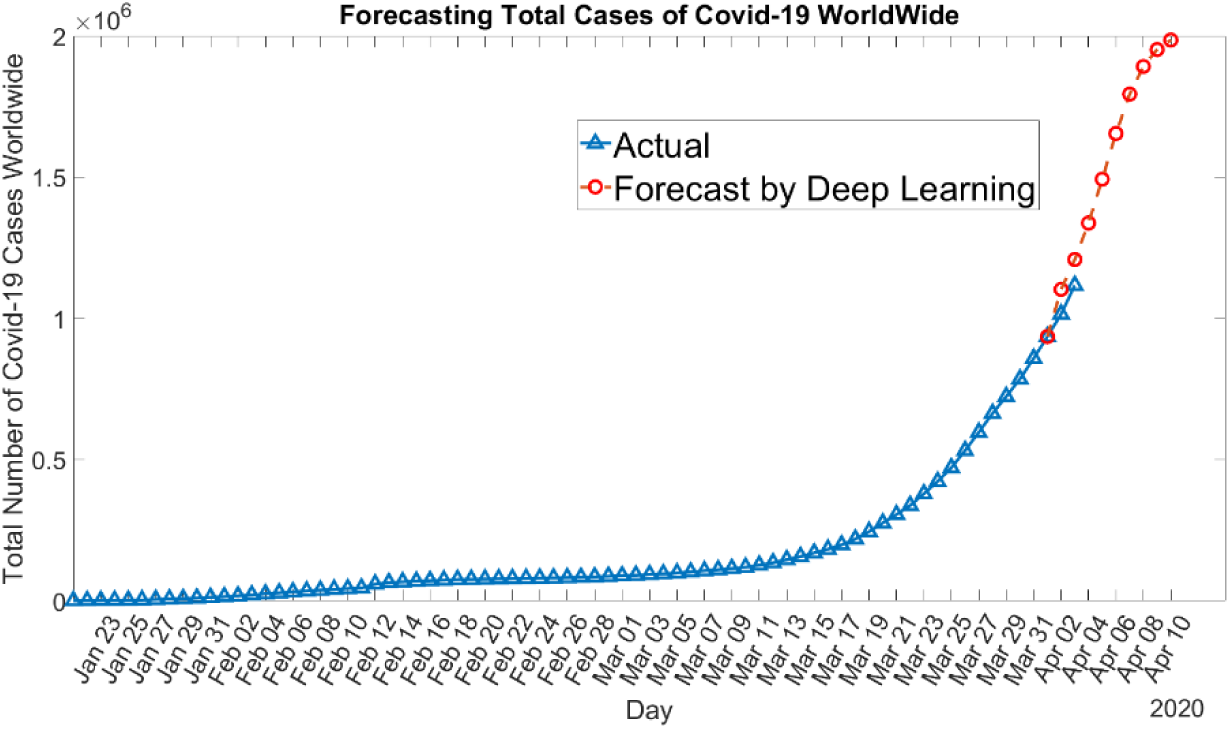
Forecasting cumulative total number of Covid-19 cases worldwide using a model with RMSE of 30258 predicts that outbreak size may reach to 2,000,000 within the next 10 days and continue to grow linearly.

**Figure 4:**
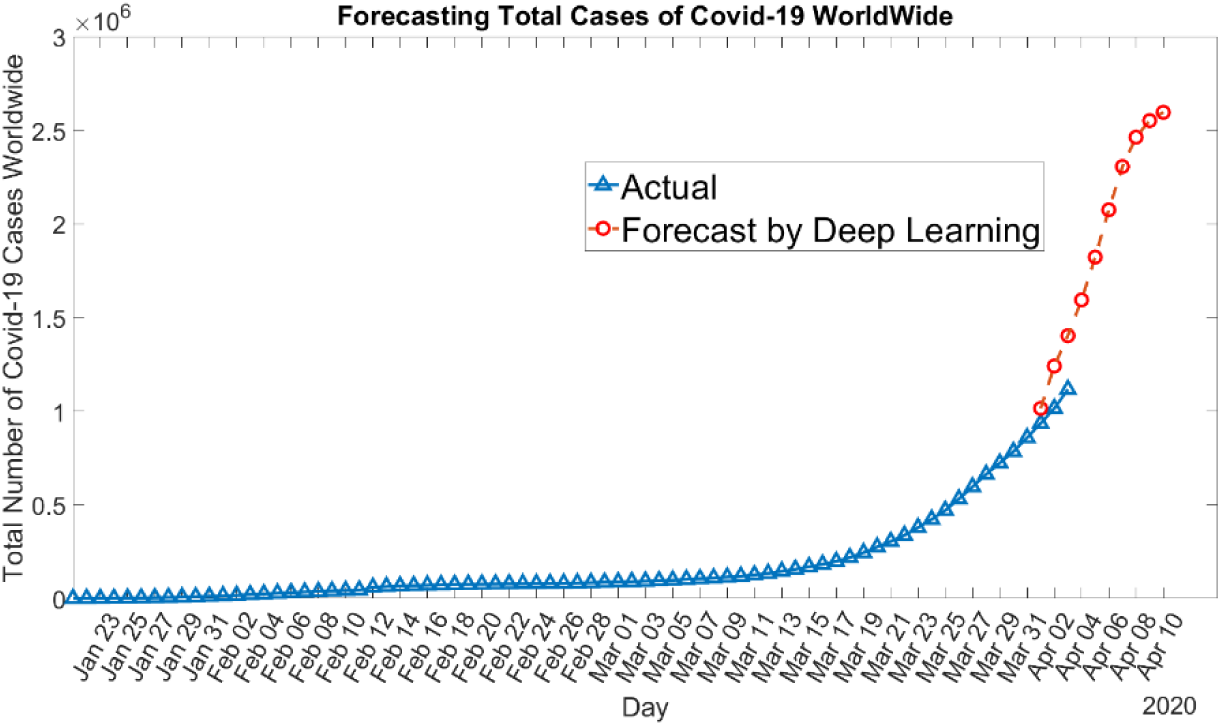
Forecasting cumulative total number of Covid-19 cases worldwide using a model with RMSE of 39699 predicts that outbreak size may reach to 2,600,000 within the next 10 days and continue to grow linearly.

**Figure 5:**
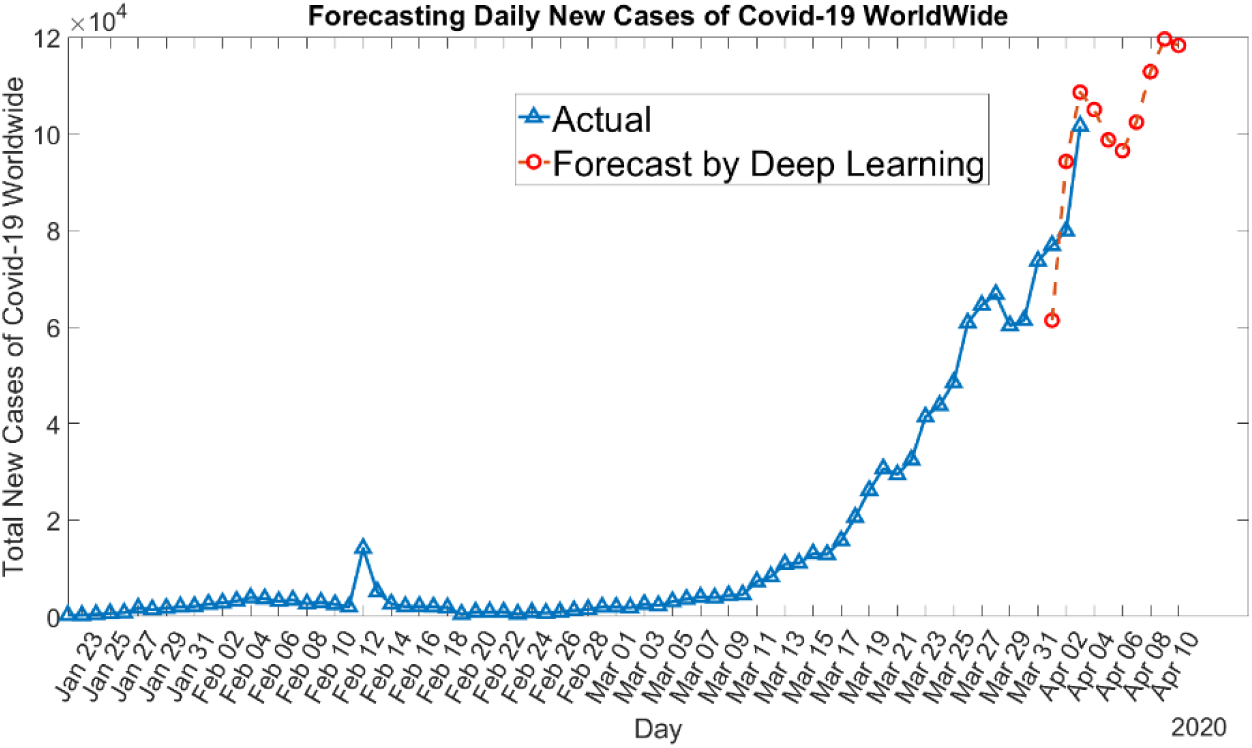
Forecasting daily new cases of Covid-19 Worldwide using a model with RMSE of 1953.5; predicts that daily new cases will be in the range of 100,000 and 120,000 within the next 10 days.

**Figure 6:**
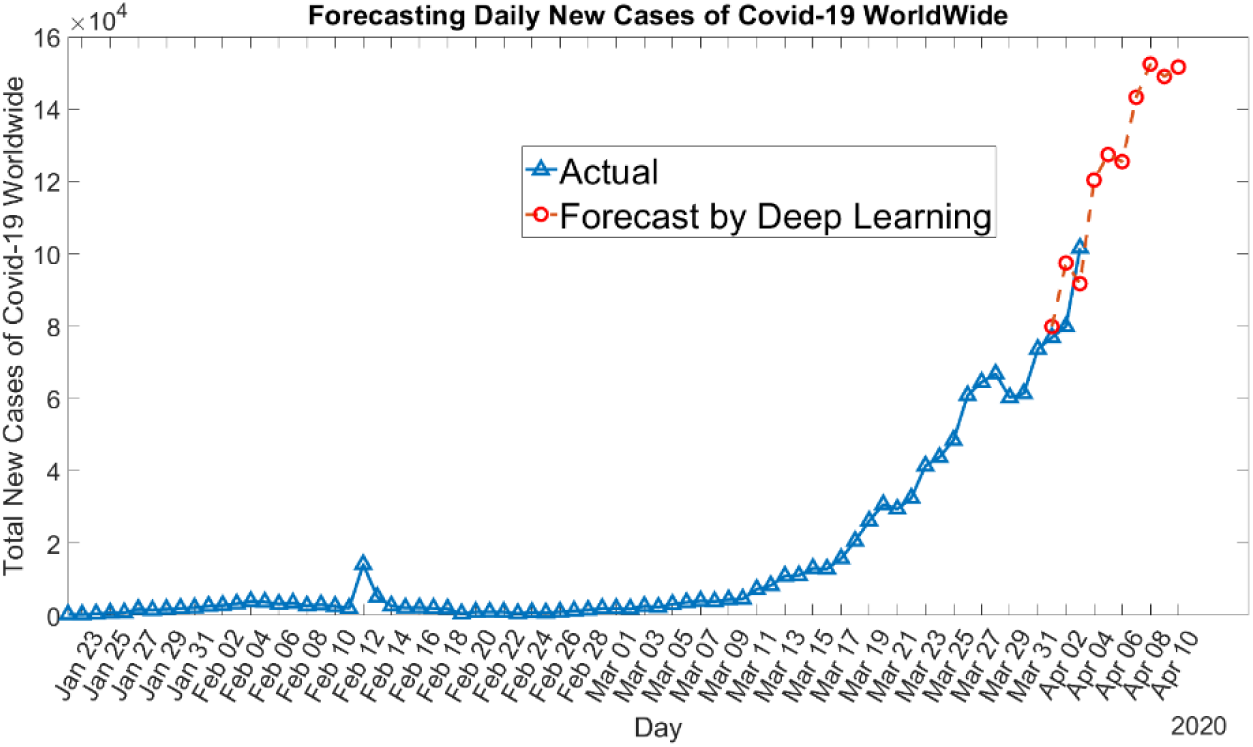
Forecasting daily new cases of Covid-19 Worldwide using a model with RMSE of 64337; predicts that might be a jump in the number of reported cases within a week and daily cases may reach to 160,000.

**Figure 7:**
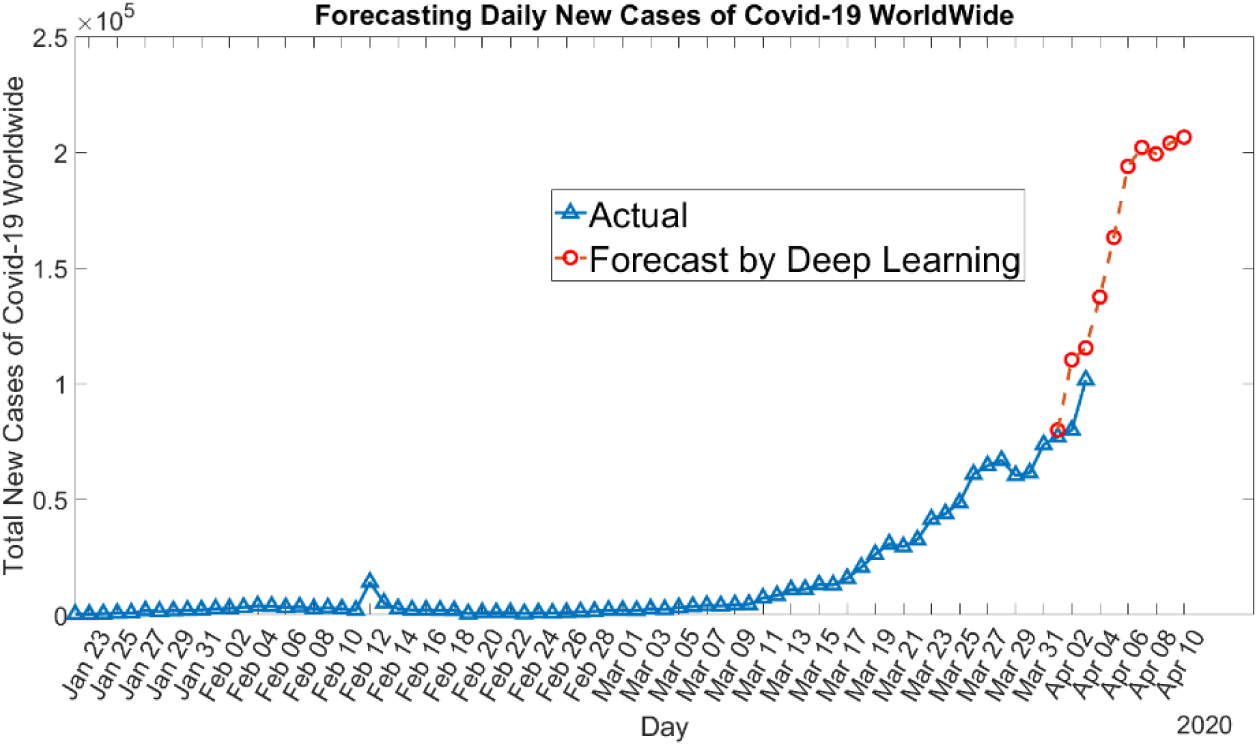
Forecasting daily new cases of Covid-19 Worldwide using a model with RMSE of 63850; predicts that daily new cases may increase to 200,000 within the next 10 days.

**Figure 8:**
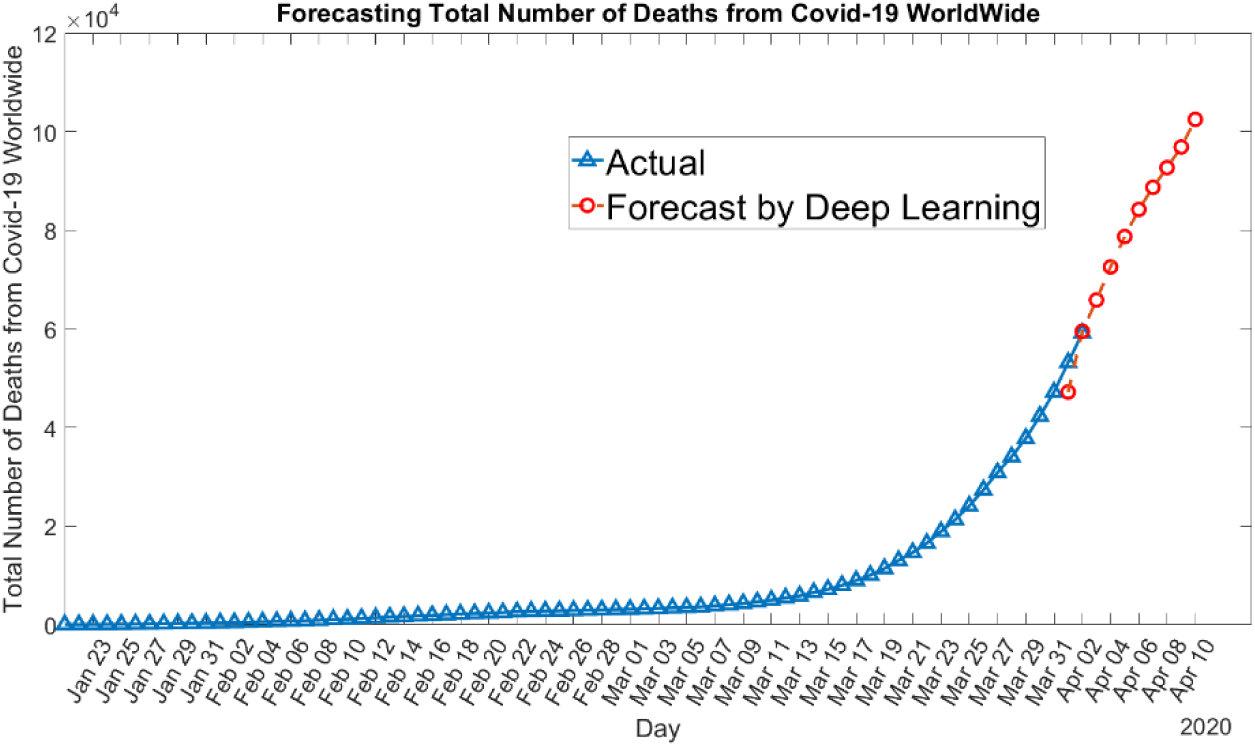
Forecasting cumulative total number of deaths from Covid-19 worldwide using a model with RMSE of 5657.1; predicts that the death toll may increase to 100,000 within the next 10 days.

**Figure 9:**
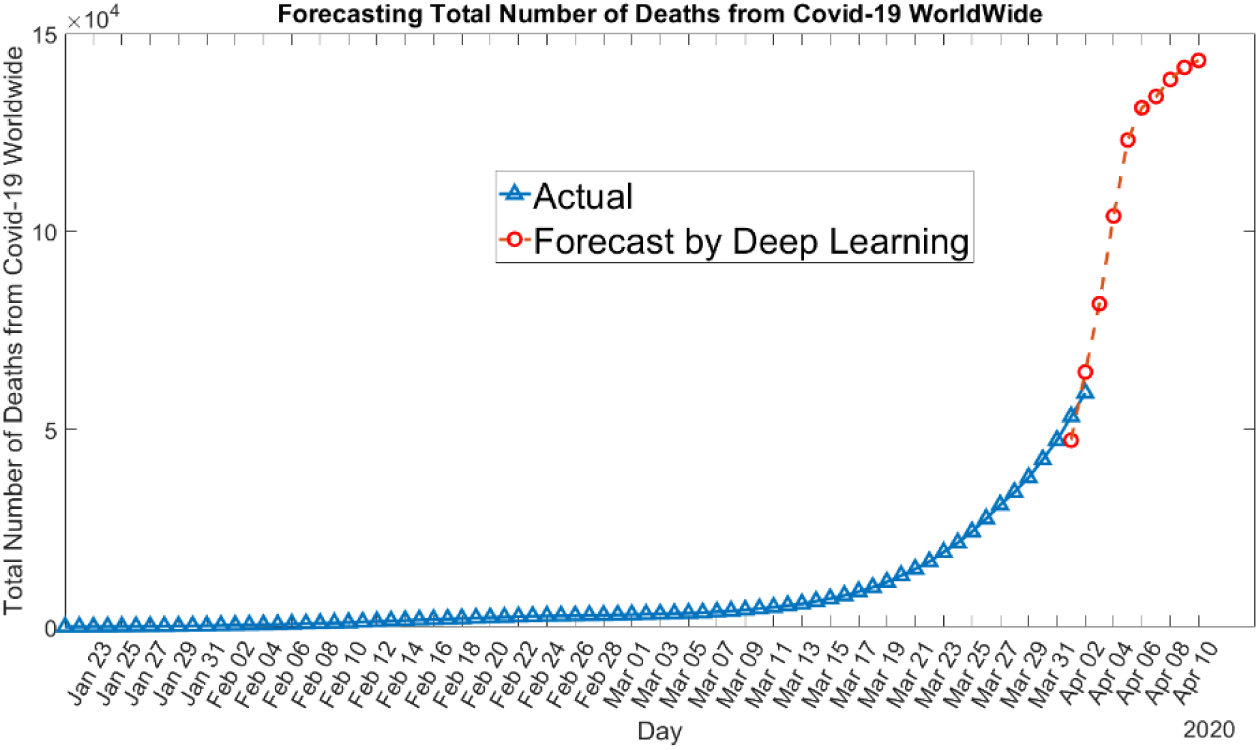
Forecasting cumulative total number of deaths from Covid-19 worldwide using a model with RMSE of 7703.3; predicts that the death toll may increase to 150,000 within the next 10 days.

### 4.3 Results and Discussions for China Forecasts

The deep learning approach provides various scenarios in China. For daily new cases, we present three different scenarios: (1) As shown in Figure 10, outbreak of the disease will nearly stop (RMSE=107.2), (2) daily new cases may increase slightly as shown in Figure 11 (RMSE=127.3) and (3) as illustrated in Figure 12, the number of new infections may increase again (RMSE=204.8). Forecasts suggest that the number of deaths in China will not increase considerably. Figure 13 shows a forecast that the number of deaths is stabilizing (RMSE=15.4).

**Figure 10:**
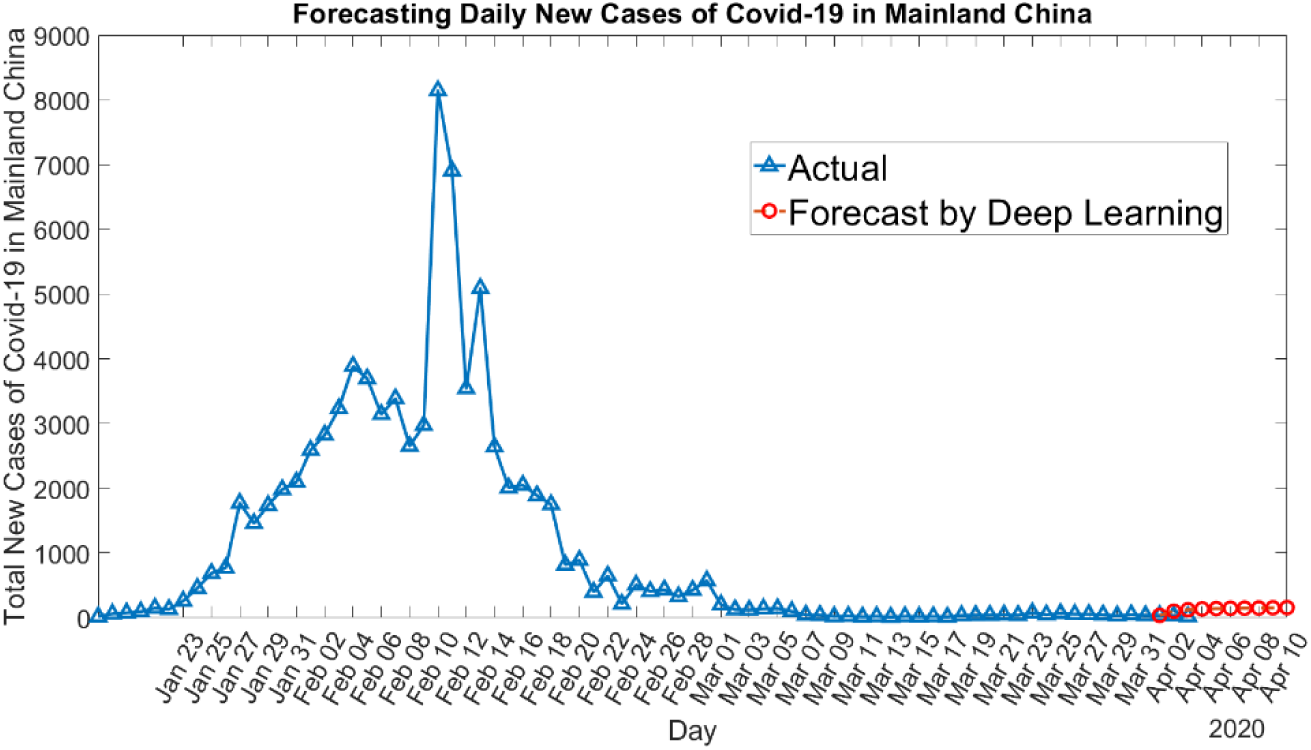
Forecasting daily new cases of Covid-19 in Mainland China using a model with RMSE of 107.2; predicts that outbreak will nearly stop

**Figure 11:**
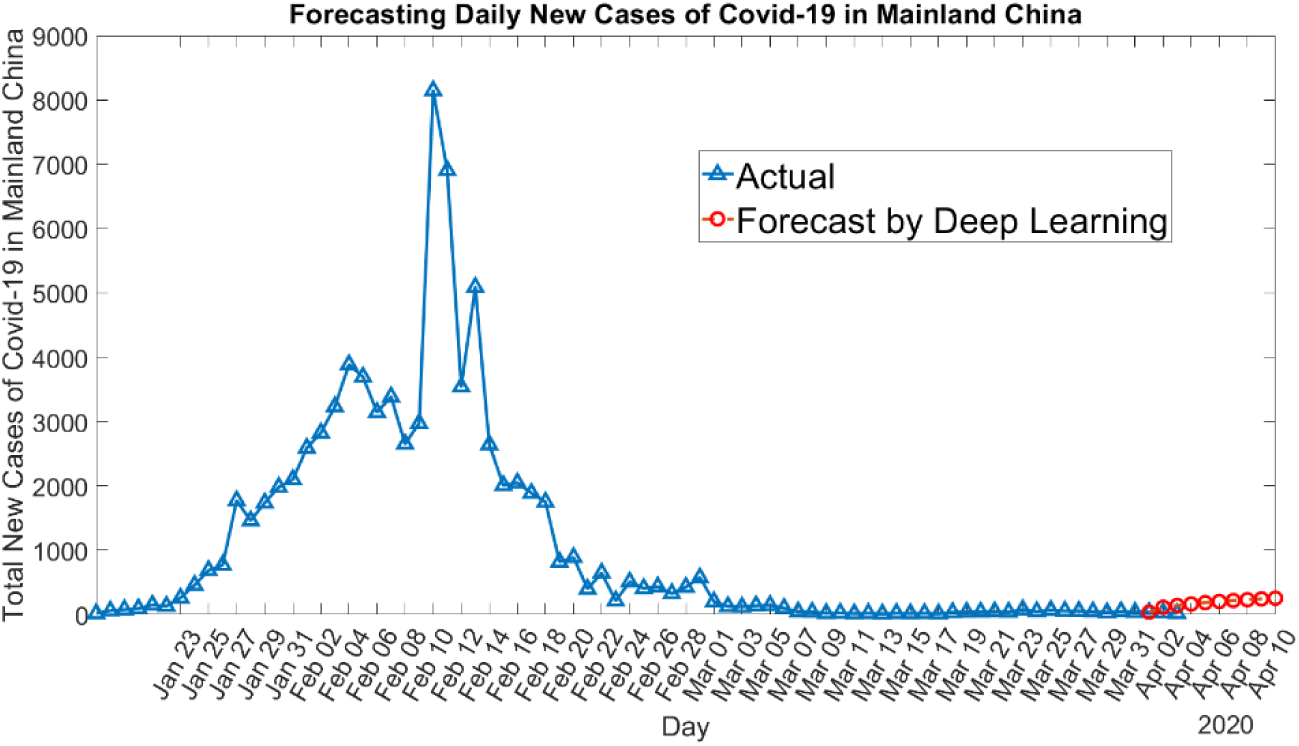
Forecasting daily new cases of Covid-19 in Mainland China using a model with RMSE of 127.3; predicts that daily new cases may increase slightly

**Figure 12:**
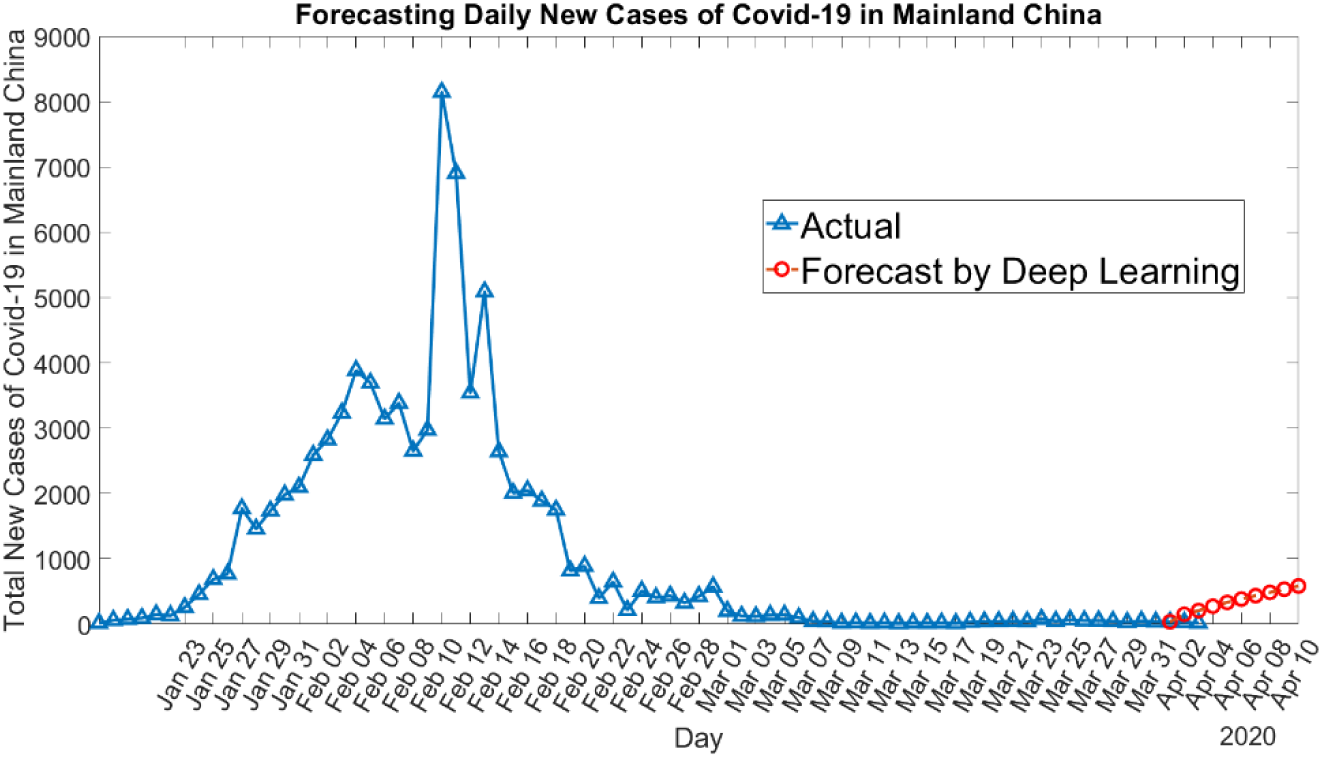
Forecasting daily new cases of Covid-19 in Mainland China using a model with RMSE of 204.8; predicts that the spread may increase again

**Figure 13.**
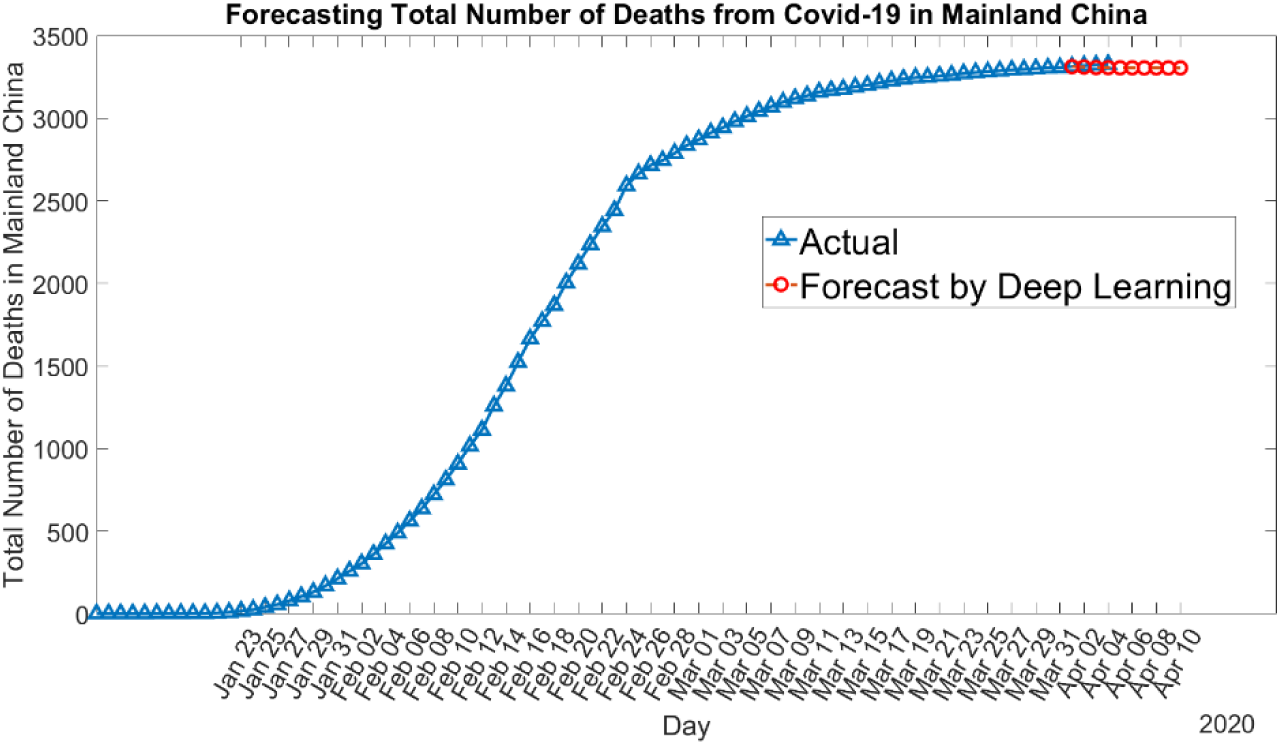
Forecasting the total number of deaths from Covid-19 in Mainland China using a model with RMSE of 15.4. predicts that the number of deaths is slowing

### 4.4 Results and Discussions for Europe and Middle East Forecasts

Comparing to China and worldwide data, we have fewer amounts of data for Europe and Middle East. However, the deep learning model forecasts considerable increase in the number of reported cases in Europe and Middle East. It forecasts that the spread will increase within the next 10 days. For Europe, the deep learning model predicts different outbreak scenarios: (1) The outbreak size may reach to 800,000 (RMSE=29139) (see Figure 14) within the next 10 days. (2) The outbreak size may reach to 1,000,000 (RMSE=59726) (see Figure 15) within the next 10 days. (3) The outbreak size in Europe may even reach to 1,800,000 within the next 10 days (RMSE=12889) as shown in Figure 16. For Middle East region, again deep learning model forecasts different outbreak scenarios: (1) The outbreak size in Middle East may increase to 90,000 as shown in Figure 17 (RMSE=6629.9). (2) The outbreak may reach to 160,000 as shown in Figure 18 (RMSE=12358). (3) The outbreak may reach to even more than 200,000 as shown in Figure 19 (RMSE=10215).

**Figure 14:**
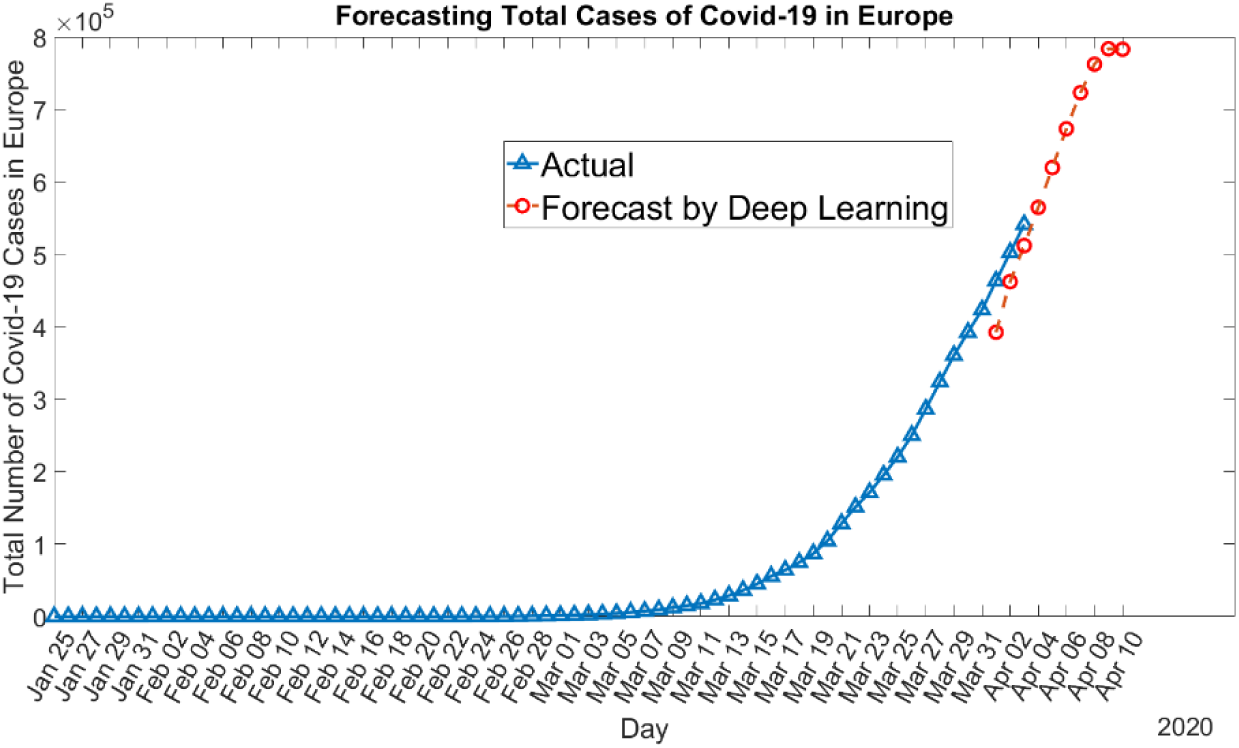
Forecasting the total number of Covid-19 cases in Europe using a model with RMSE of 29139; predicts that the spread may reach to 800,000 cases

**Figure 15:**
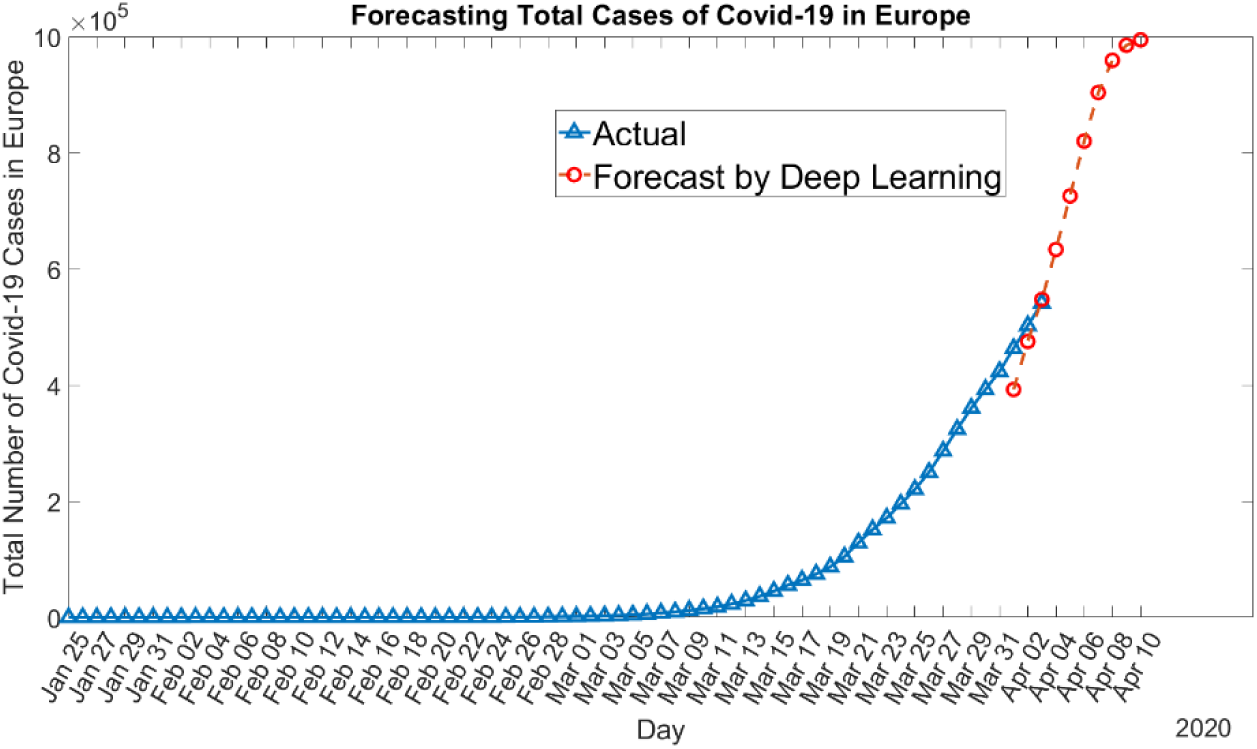
Forecasting the total number of Covid-19 cases in Europe using a model with RMSE of 59726; predicts that the spread may reach to 1,000,000 cases

**Figure 16:**
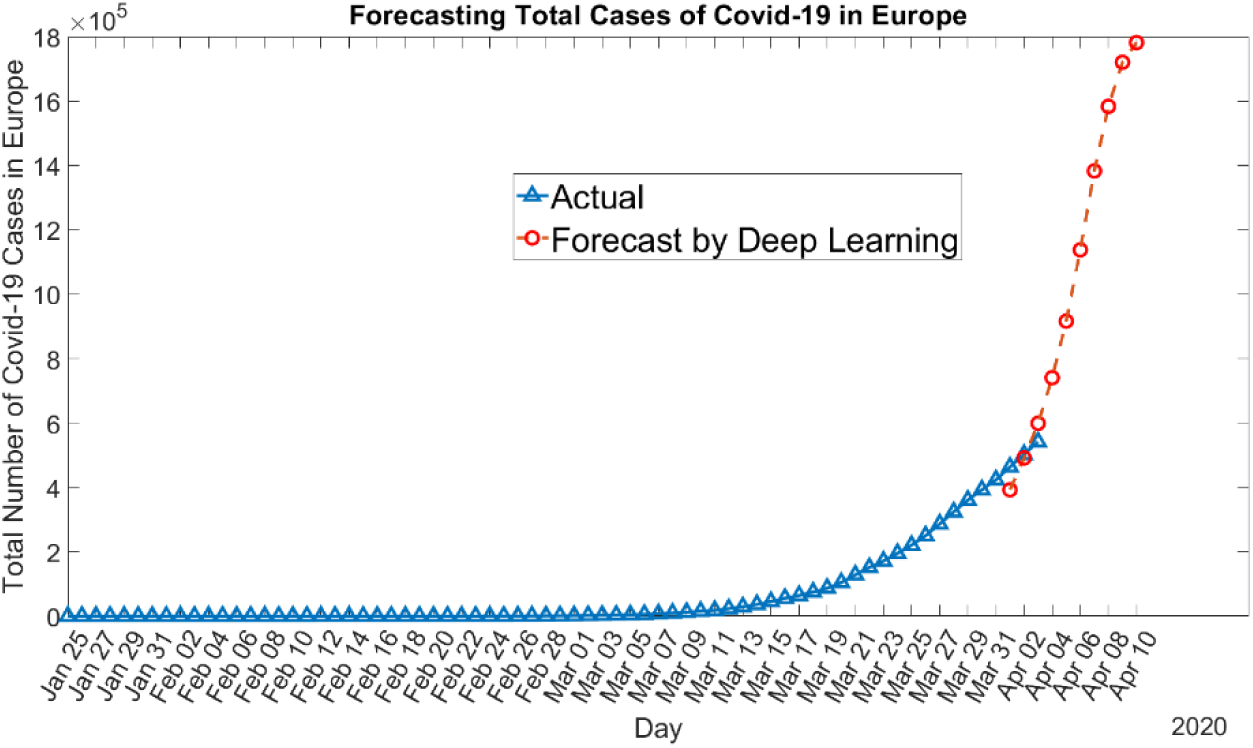
Forecasting the total number of Covid-19 cases in Europe using a model with RMSE of 12889; predicts that the spread may reach to 1,800,000 cases

**Figure 17:**
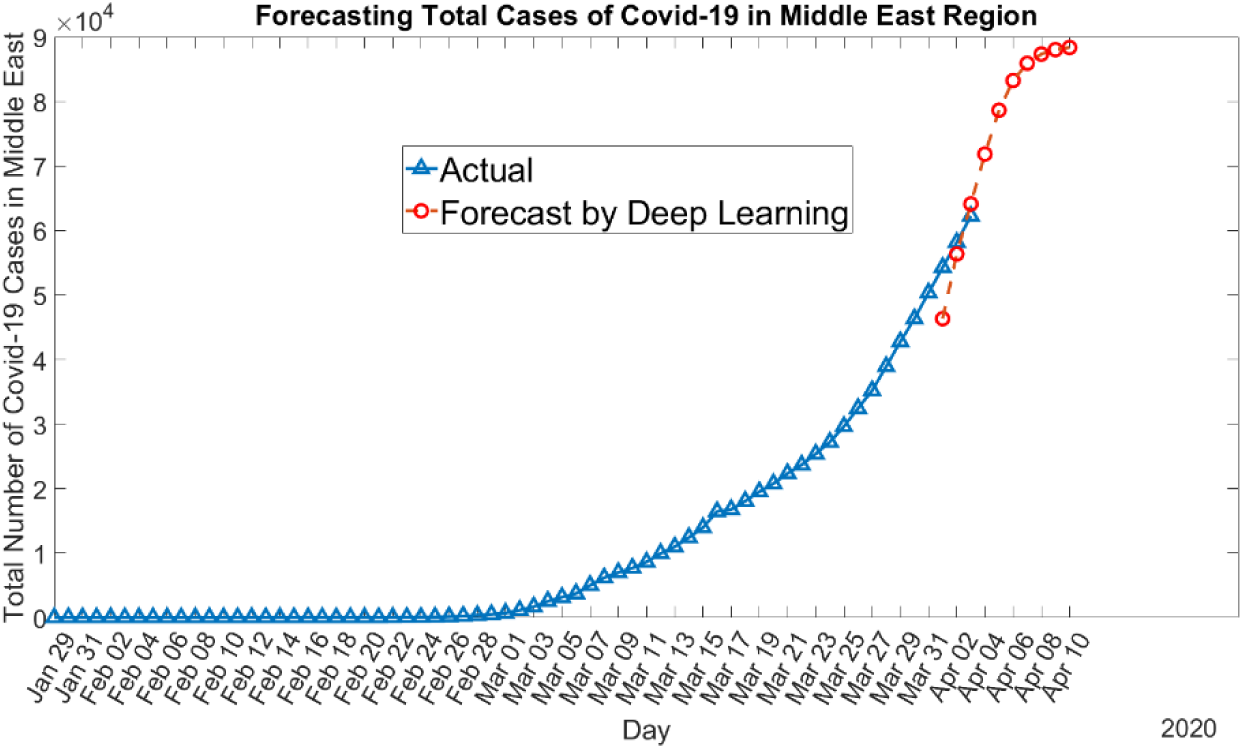
Forecasting the total number of Covid-19 cases in Middle East using a model with RMSE of 6629.9; predicts that the spread may increase to 90,000

**Figure 18:**
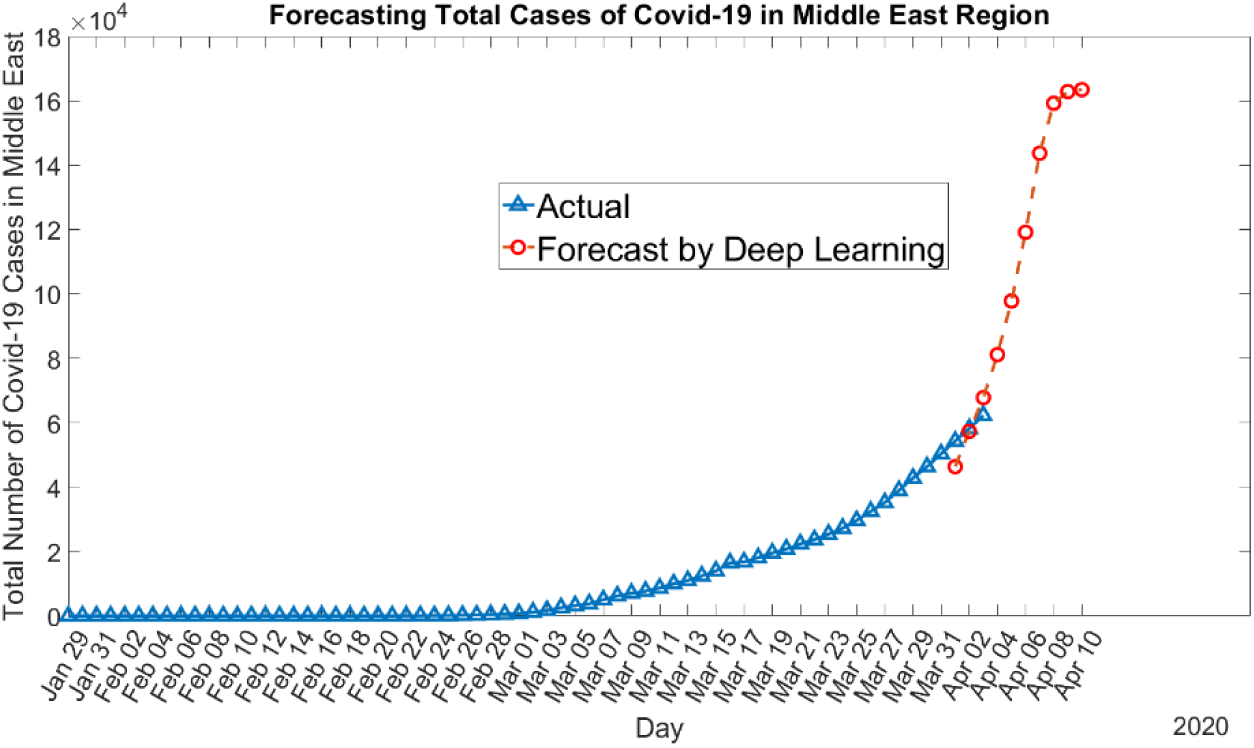
Forecasting the total number of Covid-19 cases in Middle East using a model with RMSE of 12358; predicts that the spread may increase to 160,000

**Figure 19:**
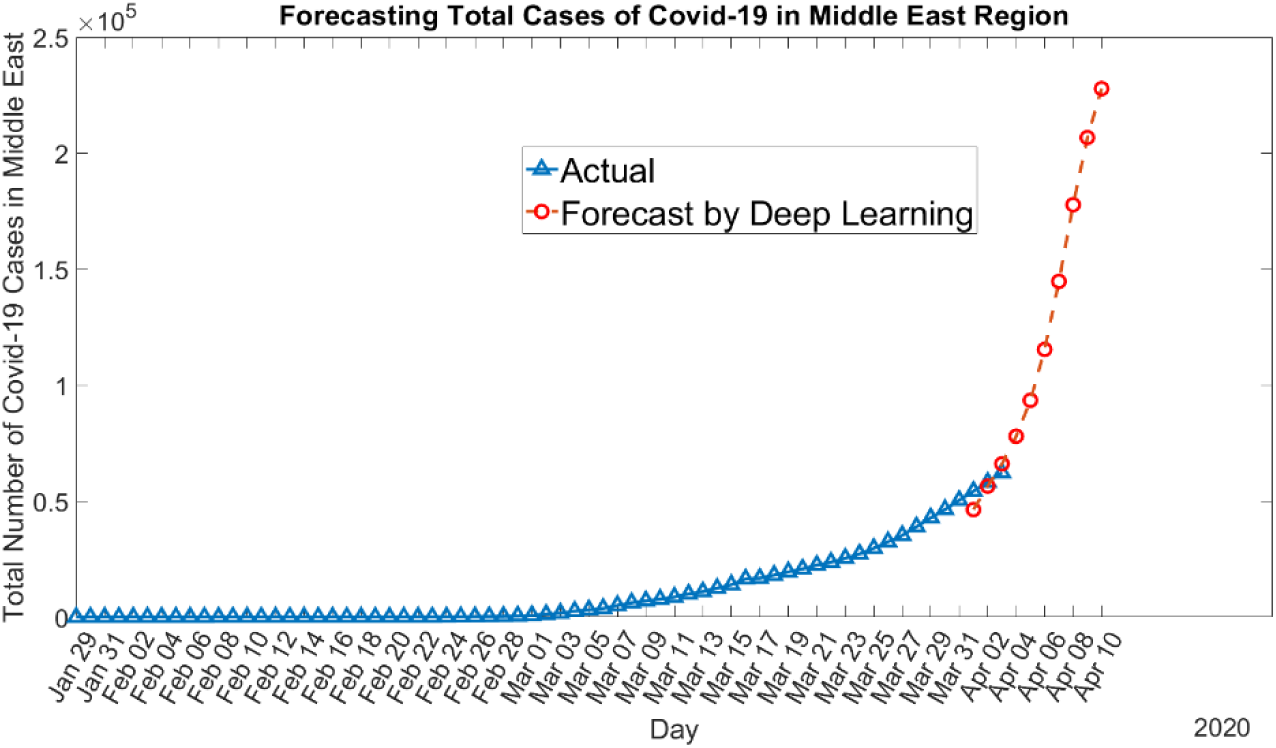
Forecasting the total number of Covid-19 cases in Middle East using a model with RMSE of 10215; predicts that the spread may reach to more than 200,000

### 4.5 Discussions

The deep learning approach provides possible scenarios and forecasts for Covid-19 regionally and worldwide. Given the forecasts, it is seen that the spread of the Covid-19 is accelerating in Europe, Middle East and worldwide. In China, we observe decrease in the number of daily reported cases, and decrease in the number of deaths. Table 3 also provides information about the expected total number of cases within the next 10 days. These findings show that the outbreak is slowing in China. While in Europe and Middle East, the number of cases is increasing considerably.

**Table 3:** Analysis of forecasts

|  | **31 March 2020**<br>**(Now)** | **10 April 2020**<br>**(Forecasted)** | **RMSE Forecast for the**<br>**last 3 known days**<br>**(7, 8 and 9 March 2020)** |
| --- | --- | --- | --- |
| Total cases of Covid-19 worldwide (scenario 1) | 858,361 | ~2,000,000 | 30258 |
| Total cases of Covid-19 worldwide (scenario 2) | 858,361 | ~2,600,000 | 39699 |
| Total deaths from Covid-19 worldwide (scenario 1) | 42,309 | ~100,000 | 5657.1 |
| Total deaths from Covid-19 worldwide (scenario 2) | 42,309 | ~150,000 | 7703.3 |
| Total Cases in Europe (scenario 1) | 423,443 | ~800,000 | 29139 |
| Total Cases in Europe (scenario 2) | 423,443 | ~1,000,000 | 59726 |
| Total Cases in Europe (scenario 3) | 423,443 | ~1,800,000 | 12889 |
| Total Cases in Middle East (scenario 1) | 50,349 | ~90,000 | 6629.9 |
| Total Cases in Middle East (scenario 2) | 50,349 | ~160,000 | 12358 |
| Total Cases in Middle East (scenario 3) | 50,349 | ~225,000 | 10215 |

## 5. Conclusions

We design a deep learning model to forecast the spread of the novel coronavirus, Covid-19, in China, Europe, Middle East region and worldwide. In this paper, we present results for the prediction of cumulative number of cases, cumulative number of deaths and daily new cases worldwide. Similarly results for Europe, Middle East and Mainland China are provided. Based on the actual time series data of Covid-19, we employ a deep neural network to predict the next 10 days. This is the first time that a deep learning model is proposed for forecasting spread of Covid-19 solely using time series data of reported cases. We provide possible scenarios for China, Europe, Middle East region and worldwide. We evaluate networks using the Root Mean Square Error (RMSE) metric. For predictions, we use networks that give the minimum RMSE on the last 3 days of actual data. The proposed model can be updated by retraining with new daily data. It can learn the patterns of data and can provide possible scenarios for future. Overall, worldwide the spread is accelerating except China. Therefore, immediate and consistent action is required by countries, authorities and individuals in order to contain the virus and stop spreading of it.

## Data Availability

We have used the publicly available data about Covid-19 statistics from World Health Organization and World-o-Meter.

